# Pre-referral stabilization practices and associated early clinical outcomes among critically ill under-five children at regional referral hospitals in Dar es salaam, Tanzania

**DOI:** 10.64898/2026.01.11.26343875

**Authors:** Nasreen Ayoub Daud, Florence Salvatory Kalabamu, Maulid Rashid Fataki, Zahra Morawej, Deogratius Mathew Kallanga, Ansaar Ismail Sangey, Rhobi Gabriel Sisa

**Affiliations:** Dept of Paediatrics/Child Health – Kairuki University Dar es Salaam, Tanzania; Dept of Psychiatry & Behavioural Medicine– Kairuki University Dar es Salaam, Tanzania; Dept of Emergency Medicine – Temeke Regional referral hospital, Dar es Salaam, Tanzania

**Keywords:** Pre-referral stabilisation, children

## Abstract

**Background:** Pre-referral stabilization is a critical yet neglected component of pediatric emergency care in low-resource settings. It involves delivering life-saving interventions— such as airway management, oxygen therapy, fluids, glucose, and anticonvulsants at the initial point of care before referral. The WHO’s ETAT Guidelines emphasize the prompt recognition and management of life-threatening conditions in children to reduce mortality. Despite Tanzania’s adoption of ETAT, the extent to which pre-referral stabilization is practiced remains poorly documented. Strengthening pre-referral care through adherence to ETAT principles is essential to improve child survival and enhance the overall emergency response system.

**Objective:** To assess the pre-referral stabilization practices and associated early clinical outcomes among critically ill children under five years referred to Amana, Mwananyamala and Temeke Regional Hospitals in Dar es Salaam, Tanzania, during the study period.

**Methodology:** A hospital-based cross-sectional study that will be conducted from January 2026 to March 2026 at Amana, Mwananyamala, and Temeke Regional Referral Hospitals in Dar es Salaam, Tanzania. Consecutive eligible under-five children (0–59 months) referred from lower-level facilities who meet WHO ETAT emergency criteria will be enrolled (target n = 420). Data will be collected using a structured, pre-tested case report form capturing socio-demographic characteristics, pre-referral stabilization measures, and early outcomes within 72 hours post-admission. The primary outcome variable will be child final status (alive vs dead). Data will be analyzed in SPSS v23 using descriptive statistics, chi-square tests, and multivariable logistic regression to examine associations between pre-referral practices and early clinical outcomes.

## INTRODUCTION

Under-five mortality remains a global health concern, particularly in low- and middle-income countries (1, 2). Although significant progress has been made in reducing child deaths, survival among critically ill under-five children continues to depend heavily on the quality of emergency care provided at the first point of contact. In resource-limited settings, delays in identifying and stabilizing severely ill children before referral often lead to preventable deterioration and death (2, 3).

Pre-referral stabilization is an essential component of pediatric emergency care that ensures critically ill under-five children receive immediate life-saving interventions before being transferred to a higher-level health facility (4). It involves identifying emergency signs and promptly initiating essential measures with the aim of correcting life-threatening physiological disturbances and preventing further deterioration during transport. Evidence from low- and middle-income countries demonstrates that many deaths among referred children occur within the first 24 to 48 hours of hospital admission, often due to delays or omissions in these early interventions at referring facilities(5-6).

One of the most important, yet often overlooked, aspects of pediatric emergency management is pre-referral stabilization. This involves the initial clinical interventions performed at the first point of contact before transferring a critically ill child to a higher-level facility. Recognizing its significance in improving child survival, the World Health Organization (WHO) has included pre-referral stabilization as a key part of the Emergency Triage Assessment and Treatment (ETAT) strategy. These actions typically include clearing and maintaining the airway, providing oxygen for respiratory distress, controlling seizures, administering glucose in hypoglycemia, starting appropriate antibiotics for suspected infection, and initiating fluid resuscitation in shock. The goal of these actions is to stabilize vital functions, enabling either safer transfer or in-facility care as appropriate(7).

In the Tanzanian context, regional referral hospitals such as Amana, Mwananyamala, and Temeke function as major referral points within the urban health system. These hospitals regularly receive critically ill children referred from surrounding health centers and dispensaries. Although Tanzania has adopted ETAT nationally as a standard for emergency pediatric care, the extent to which pre-referral stabilization is being implemented in practice prior to transfer remains insufficiently documented, particularly in high-volume urban areas like Dar es Salaam, Tanzania (8).

Despite Tanzania’s national adoption of the WHO ETAT guidelines, under-five mortality remains alarmingly high at 43 deaths per 1,000 live births, with up to 75% of these deaths occurring within the first 24–48 hours of hospital admission (9). Many of these deaths involve children presenting with preventable and treatable conditions such as pneumonia, sepsis, malaria, and severe dehydration (9). These critically ill children are frequently referred from lower-level health facilities to regional referral hospitals; however, it is unclear whether they receive appropriate emergency stabilization prior to transfer, including interventions such as oxygen therapy, intravenous fluids, and first-dose antibiotics as recommended by WHO (10).

In Tanzania, most studies have concentrated on neonates or adult trauma cases, highlighting gaps in pediatric emergency care. However, peer-reviewed research that systematically examines pre-referral stabilization of under-five children in regional referral hospitals using WHO ETAT criteria is still scarce. No analytic study from Tanzania has been published to date, although related theses, dissertations, or other forms of grey literature may exist (11, 12). Furthermore, little is known about how these practices or their absence impact early clinical outcomes such as mortality, deterioration on arrival, or intensive care admission, particularly in high-volume urban referral hospitals. A tertiary-level study identified systemic deficiencies, including delayed recognition, poor referral documentation, and a lack of essential resuscitation supplies (12). Whether such gaps are also present at regional referral hospitals in Dar es Salaam remains unknown. This lack of data limits the ability to identify weaknesses in the referral system, implement targeted quality improvements, and may contribute to 15–20% of otherwise preventable child deaths (12). There is an urgent need to evaluate pre-referral stabilization practices and their association with early outcomes to inform data-driven strategies for improving pediatric emergency care.

## METHODS

### Study design

This will be a hospital-based cross-sectional study conducted at Amana, Mwananyamala, and Temeke Regional Referral Hospitals.

### Study area

The study will be conducted at Amana, Mwananyamala and Temeke Regional Referral Hospitals located in Dar es Salaam, Tanzania. These are three regional hospitals in the districts of Ilala, Kinondoni and Temeke, respectively. Each hospital has pediatric emergency and neonatal care units that routinely receive a high number of referrals of critically ill children under five from lower-level health facilities across the city. These hospitals were selected because they represent all three districts of Dar es Salaam and are the main referral centers for pediatric emergencies in the region.

### Target and study population

The target population will be the critically ill children under five years of age attending Amana, Mwananyamala and Temeke Regional Referral Hospitals. The study population will consist of all critically ill children.

### Sampling methods

The three Regional Referral Hospitals in Dar es Salaam were purposively selected as study sites because they serve large number of children with diverse illnesses and have a high volume of admissions. The study will employ a consecutive sampling technique, where all eligible under-five children who meet the inclusion criteria and whose mothers/caregivers provide informed consent will be consecutively enrolled until the required sample size is reached. Consecutive inclusion is used to minimize selection bias, ensure the sample reflects real clinical flow and case-mix in high-volume emergency and neonatal units, and maintain feasibility within a time-limited study period

### Sample size estimation

The sample size for the study was calculated using the following Cochran formula 1963:

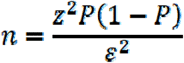

Whereby:

n = Minimum required sample size

Z = Percentage point of the normal distribution corresponding to the level of confidence. If the level of significance is 95% then Z is 1.96

□ = Maximum likely error/margin of error, i.e. 0.05

P = Prevalence of pre-stabilized critically ill (56.6%) using the findings of Mpokigwa and colleagues among children admitted to Muhimbili (2022).

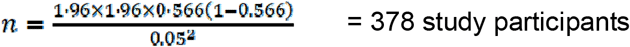

Adjusting for a 10 % non-response, the required sample size will be 420 study participants.

#### 3.3.2 Proportional allocation of study participants

To ensure a representative allocation of study participants across the three Hospitals, sample size distribution will be based on average monthly admissions to EMD statistics obtained from hospital records over the past year 2024, as shown in Table 1 below.

**Table 1:**
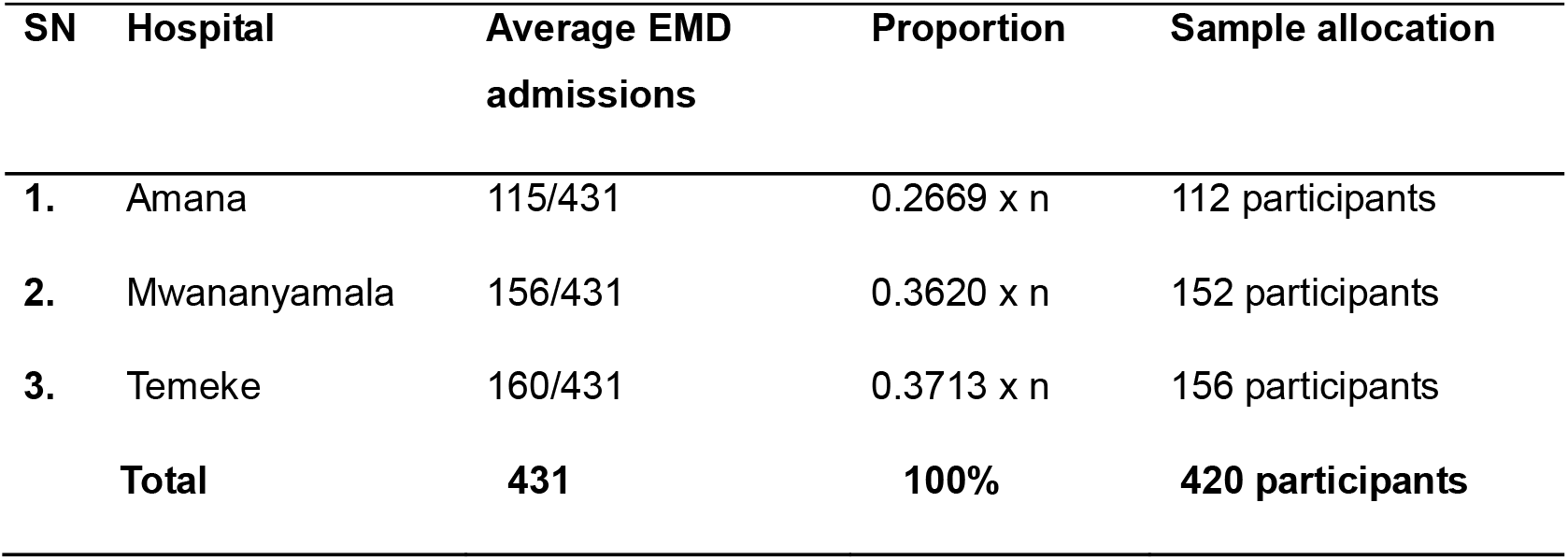
Allocation of study participants to each Hospital.

| SN | Hospital | Average EMD admissions | Proportion | Sample allocation |
| --- | --- | --- | --- | --- |
| 1. | Amana | 115/431 | 0.2669 x n | 112 participants |
| 2. | Mwananyamala | 156/431 | 0.3620 x n | 152 participants |
| 3. | Temeke | 160/431 | 0.3713 x n | 156 participants |
| Total |  | 431 | 100% | 420 participants |

### Procedures for Data Collection

#### Data collection tools

The main data collection tool will be a case report form (CRF) to collect information from each participant enrolled in the study.

It will be used to assess pre-referral stabilization status, magnitude, and their association with clinical outcomes among pediatric patients below 5 years referred to Amana, Temeke, and Mwananyamala Hospitals. The tool will have three separate sections:

**Part I**: Socio-demographic and anthropometric data of study participants and the parents/caretakers, like date of birth (age), child’s sex, caregivers’ employment status, caregivers’ level of education, and caregivers’ marital status.

**Part II:** Pre-referral information: level of the primary facility, reason for referral, treatment before referral, transit time from primary health facility to secondary health facility, Pre-referral stabilization (i.e. stabilized vs. not stabilised).

**Part III:** Early clinical outcome up to seventy-two hours: Alive (stable, unstable) vs dead

## Data collection methods

Each day, the PI and trained research assistants will visit the Pediatric Emergency Unit and Neonatal Unit at Amana, Mwananyamala, and Temeke Regional Referral Hospitals to identify eligible referred children. After initial clinical stabilization, written informed consent will be obtained.

Baseline (t_₀_): demographics, referral source, time to referral, triage category, vital signs, and pre-referral stabilization actions will be abstracted from referral notes and admission notes. Prospective follow-up: standardized chart reviews will be conducted at baseline (t_o_), 12, 24,36,48, 60 and 72 hours post-admission to record early outcomes (alive/death) and key in-hospital stabilization/process measures (oxygen in use, IV/IO patency, glucose checks, first-dose antibiotic timing). The design is appropriate for collecting information on specified time-related intervals of critical care aspects and adherence to established protocols.

## Eligibility criteria

### Inclusion criteria

- Children aged 0–59 months of age
- The child is in a critical ill state
- The child must have been referred from the lower facilities

### Exclusion criteria

- death on arrival
- immediate referral on arrival
- Children in need of immediate life-saving interventions (intubation, cardiopulmonary resuscitation)
- If the child was previously enrolled in the study during the same illness and same center.

## Withdrawal criteria

Participants will be withdrawn from active follow-up under any of the following conditions:

1. Consent withdrawn by the parent/guardian at any time,
2. Lost to follow-up.

Once withdrawn, no further data will be collected beyond the point of withdrawal. This ensures that participation remains voluntary and that data integrity and ethical principles are maintained.

## Study variables

### Dependent Variable

The dependent variable will be the 72-hour clinical outcome, categorized as follows:

- 1. Alive
- 2. Dead

### Main Independent Variable (Exposure)

The main independent variable will be the pre-referral stabilization status at admission, defined according to the performance of key WHO-ETAT stabilization interventions. It will be categorized as:

- Adequately stabilized (Only if the child is alert and has (in totality – not singly) between 90% - 100% Oxygen saturation, pule rates of 80-160 b/min, respiratory rate of 22-60 b/min, axillary temperature of 36 degrees Celsius to 37.5 degrees Celsius).
- Inadequately stabilized (A child who has derangement in any of the following conditions above)

### Independent variables

All numerical variables will be collected in their raw continuous form without any categorisation at the data collection stage. Any categorisation, if required, will only be done later during data analysis.

Independent variables: will include Child’s age – continuous variable, parents’/caretakers’ age – continuous variable in completed years, child’s weight – continuous variable, time of birth – continuous variable, and vital signs of the child (oxygen saturation – continuous variable, pulse rate – continuous variable, respiratory rate – continuous variable).

Child’s gender – dichotomized into male-1 versus female-2 (categorical variable), parents’/caretakers’ employment status – categorical variable, parents’/caretakers’ level of education – categorical variable, parents’/caretakers’ marital status – categorical variable, parents’/caretakers’ employment status – categorical variable, level of referring facility – categorical variable, number of siblings – discrete variable, AVPU – discrete variable, and pre-referral stabilisation status (categorical, binary – 1 = stabilised, 2 = not stabilised).

## Ethical considerations

The study will adhere strictly to ethical principles to ensure respect, protection, and fairness for all participants, under national and international research ethics guidelines.

The study involves minimal risk to participants, as no additional interventions will be introduced outside routine care. Data will be collected through observation and review of patient records after written informed consent has been obtained. Participation will be entirely voluntary, and refusal to participate will not affect the child’s access to treatment. The study will adhere to the principles of autonomy, beneficence, non-maleficence, and justice.

Informed written consent will be obtained from each child’s parent or legal guardian. For guardians who are illiterate, the consent form will be read aloud in Kiswahili and a thumbprint taken in the presence of an impartial witness. The study’s purpose, procedures, risks, benefits, and the right to withdraw at any time will be clearly explained.

To maintain confidentiality, each participant will be assigned a unique identification number. Names and other personal identifiers will not appear on any data collection forms. All paper-based data will be securely stored in locked cabinets, and electronic data will be saved in encrypted, password-protected files accessible only to the research team. Only the principal investigator and trained research assistants will access patient medical records for data extraction, and no identifying information will be recorded on study tools. Data will be used solely for research purposes and will be destroyed after the retention period in accordance with ethical guidelines.

### Provision for Medical and Psychosocial Support

All children enrolled in the study will remain under the direct care of the attending pediatric and nursing teams in their respective hospitals. The study will not interfere with clinical management. If a participant develops any complication, deterioration, or distress during the study period, the principal investigator or research assistant will immediately inform the attending clinician for prompt medical attention.

Psychosocial support, including counselling, will be provided by the hospital’s social welfare officer or the pediatric counselling unit as per existing hospital protocols. This ensures that all participants receive comprehensive medical and emotional care throughout their participation in the study.

### Risk–benefit to participants

Participants may experience indirect benefits through improved attentiveness of health workers during stabilization and documentation. Risk is minimal (confidentiality only) and mitigated via unique study IDs (no names), locked storage for paper forms, encrypted/password-protected electronic files, and restricted access. Participation is voluntary, with the right to withdraw at any time without impact on treatment.

## Benefits of the study

By showing where gaps exist in how critically ill children are stabilized before referral, the findings will help doctors, nurses, and hospital leaders improve the quality of emergency care. The results will support more focused ETAT training, stronger referral systems, and better access to essential emergency supplies.

Although the study does not provide direct therapeutic benefit to participants, it indirectly benefits both the children enrolled and the wider community through improved stabilization practices, better preparedness of health facilities, and faster, safer referrals. In the long term, these improvements can strengthen emergency care capacity and reduce preventable illness and deaths among under-five children.

### Protection of Vulnerable Participants

This study involves critically ill under-five children, who are considered a vulnerable group. To ensure their safety, children requiring immediate life-saving interventions such as intubation or cardiopulmonary resuscitation will not be enrolled in the study. For those who will be enrolled in the study, Informed consent will be obtained from parents or guardians only after the child has been clinically stabilized, and no study procedure will delay or interfere with emergency treatment. The study is purely observational and will not alter the standard clinical management provided by the treating team.

### Ethical clearance

Ethical approval for the study will be sought and obtained from the Institutional Research Ethics Committee (IREC) of Kairuki University. Permission to carry out the study will be requested from the Administration of the three Regional Referral Hospitals in Dar es Salaam.

### Reliability and validity of study tools

The pre-designed tool will be modified and reviewed by experts prior to a test at another private Hospital. Specifically, reliability will be assessed using test-retest analysis. Reliability analysis will be summarised using 4th Guttman’s index as well as Cronbach’s alpha coefficient. Moreover, content validation will form the mainstay of validity assessment. A minimum sample of (15% of the total minimum sample size), 63 children will be used for tool validation at a private hospital in Dar es Salaam.

## Study limitations

While the study is expected to provide valuable insights into adherence to the WHO ETAT pre-referral stabilization Guidelines and associated clinical outcomes, there are potential limitations. One major concern is the loss to follow-up, as participants will be monitored for up to seventy-two hours after admission. Sources of loss of follow-up may include communication breakdown as well as premature discharge from the wards (e.g. due to failure to pay hospital costs/treatments costs). Moreover, children who will be referred to the tertiary hospital Muhimbili National Hospital, as well as those patients arriving at the study centre on a self-referral basis, may not be readily available and therefore, likely underestimate the true burden of pre-referral stabilization.

Another limitation is the likelihood of recall bias from the parents/guardians. To mitigate this, the study will rely on triangulation using multiple data sources, such as clinical charts, referral documentation, and direct observation. Moreover, being an observational study, it will not be possible to establish causality between inadequate stabilization and clinical outcomes. The findings will be limited to associations rather than cause-and-effect conclusions.

### Data management plan

The PI will securely store paper-based case report forms (CRFs) in locked cabinets accessible only to authorized personnel. Soft copy of the data will be saved in a password-protected protected hard-drive only known to the principal investigator. Data will be made available upon reasonable request from the principal investigator, upholding the confidentiality and anonymity of the participants at all times.

### Study procedures in the field

Prior to data collection, six Research Assistants - Medical Officers, Registered Nurses or Clinical Officers at Emergency Departments at the three Regional Referral Hospitals will receive comprehensive training on the study protocol – how to screen for eligibility criteria; to request for informed consent; to undertake data collection by conducting interviews with caretakers guided by the Case Report Form and also review medical records to retrieve pertinent information as per referral checklists.

### Data coding and cleaning

To ensure the accuracy, consistency, and reliability of data collected, verification will begin at the point of collection. Research Assistants will conduct immediate checks for completeness and clarity of each Case Report Form (CRF) after it has been filled, resolving any missing data.

To further enhance data accuracy, the Principal Investigator will conduct routine spot checks and audits to ensure adherence to the established data collection protocols.

The electronic data management system will maintain audit trails to log all changes, ensuring accountability and transparency in data handling.

The study will ensure the confidentiality of study participants’ data by assigning unique study participant identification codes in place of personal identifiers. The Study team will encrypt all electronic data, and physical access to documents will be controlled. The study team will use multi-factor authentication for accessing sensitive digital files. The study team will retain data necessary to meet the study objectives and will then dispose/retain it as per the Institutional data retention policies.

## Data analysis plan

The Principal Investigator will conduct data analysis in collaboration with a Biostatistician. Immediately following the data collection exercise, and at the end of each day, data entry and subsequent exploratory and final analysis will be done using SPSS version 23 (IBM Corporation, Armonk, New York, USA). Double data entry will be done using at least 1 research assistant and the PI herself daily. For all inconsistent data, a quick re-appraisal will be done by the PI herself. Thereafter, data will be stored inside PI’s laptop until analysis time.

Initial data analysis will include exploratory data analysis, which will employ data summarization and trend checking before the main analysis process. Categorical variables (each stabilization component at recruitment, 12-, 24-,36-, 48-, 60-, and 72-hour outcome status) will be summarized as frequencies and proportions and displayed using bar charts, as well as box plots to facilitate comparison across subgroups. Continuous variables (e.g. time to referral, age) will be summarized using medians and interquartile ranges (IQRs).

Graphical and descriptive statistical tools such as histograms, line graphs and frequency tables will be used to summarise data.

The dependent variable will be the 72-hour clinical outcome (coded as 1 = alive, 2 = died). The main exposure will be adequacy of pre-referral stabilization at admission (t_₀_), defined as performance of key WHO-ETAT interventions (oxygen, IV fluids, glucose, anticonvulsant, and first-dose antibiotic). Covariates will include age, sex, diagnosis, referral source, time to referral, and hospital stata.

To determine the magnitude of pre-referral stabilization, the proportion of participants who received adequate stabilization before referral will be calculated against the total number enrolled. Non-performance of individual stabilization components (e.g., oxygen therapy, fluid resuscitation, glucose administration, first-dose antibiotics) will be summarized as percentages and presented in tables and bar charts.

To identify the most frequently omitted components of the WHO-recommended pre-referral stabilization protocol, each stabilization component (e.g., oxygen therapy, fluid resuscitation, glucose administration, anticonvulsants, and first-dose antibiotics) will be analyzed separately. Frequencies and percentages will be computed for each stabilization measure (performed vs. not performed). Findings will be summarized in tables and box plots/bar charts to illustrate the most commonly omitted elements of pre-referral stabilization across three hospitals.

To assess the association between pre-referral stabilization practices and early clinical outcomes among critically ill under-five children, the Chi-square test (or Fisher’s exact test where appropriate) will be used for bivariate analysis. Variables with a p-value ≤ 0.2 in the bivariate analysis will then be entered into a multivariable binary logistic regression model to identify independent predictors of early clinical outcome. The dependent variable will be the 72-hour clinical outcome (alive = 1, dead = 2). The main independent variable will be pre-referral stabilization status at admission (adequate vs. inadequate). Covariates such as age, sex, diagnosis, time to referral, and referral source will be included in the model. Results will be reported as Adjusted Odds Ratios (aORs) with 95% confidence intervals (CIs), and statistical significance will be defined at p < 0.05.

## Contribution of the research study

### To science

-The findings of this study will be shared as original data (via publications) on pre-referral stabilization and associated factors among the under-five-year population in Dar es Salaam hospitals, data of which has no evidence of being published in these settings before.

-No prior evidence of findings on what are the pre-referral stabilized approaches to under-fives admitted to Dar es Salaam hospitals.

### To clinical practice

- Findings of this study will bring about improvement in clinical practices via addressing findings to what is missing in standard local practice.
- Possibilities of capacity building among healthcare workers, given the gaps in knowledge and practice in pre-referral stabilization of the under-five-year population.

### To policy

- Findings of this study are likely to bring about new guidelines and formulations regarding pre-referral stabilization of under-fives admitted with emergency and priority signs at the primary healthcare level in Tanzania.

## Data Availability

This is a research protocol. No data available at this stage.

## Dissemination of study findings

The study findings will be compiled into a dissertation and submitted to the Department of Pediatrics and Child Health, Kairuki University. Copies will be deposited in the University Library and provided to the three Regional Referral Hospitals in Dar es Salaam (Amana, Mwananyamala, and Temeke).

Academic dissemination. A manuscript will be prepared for submission to a peer-reviewed journal. Findings will also be presented at scientific conferences.

Community-facing and policy dissemination. A two-page Kiswahili brief summarizing key findings and practical recommendations will be produced, posted on the Emergency Department and Pediatric ward notice boards at the three hospitals. Facility feedback meetings will be convened with clinical leadership, nursing/triage staff, and caregiver/community representatives. A concise policy brief will be prepared for District/Municipal Health Management Teams and the Ministry of Health to support integration of recommendations into local practice.

## Notes

### Competing Interest Statement

The authors have declared no competing interest.

### Funding Statement

This study did not receive any funding

### Author Declarations

Name of the institution: Kairuki University Name of IRB board: Institutional Research Ethics Committee (IREC)

